# Mentoring Matters: Evaluating The Black Physicians of Canada Mentorship Program

**DOI:** 10.1101/2024.12.07.24318652

**Authors:** Chikaodili Obetta, Anjali Menezes, Nivetha Chandran, Onaope Egbedeyi, Maryam Taghavi, Modupe Tunde-Byass, Catherine Yu, Csilla Kalocsai, Umberin Najeeb, Rukia Swaleh, Mireille Norris

## Abstract

**Purpose:** Black individuals face significant barriers in medicine, contributing to their underrepresentation as physicians and emphasizing the need for systemic change. From admissions processes to program design, medical schools often uphold outdated racial practices that disadvantage Black learners. Increased representation in medical schools benefits learners and improves care for diverse patient populations. Mentorship has proven essential in fostering success in higher education and can mitigate barriers to career progression. However, many Black learners face barriers to accessing quality mentorship despite its proven benefits in fostering equitable opportunities and career progression. The Scarborough Charter outlined 58 Canadian institutions committed to advancing Black inclusion in higher education through mentorship and accountability measures. In alignment with this goal, the Black Physicians of Canada (BPC) launched a racially concordant mentorship program. This study aimed to explore participants’ experiences and provide recommendations for future program iterations.

**Methods:** This study employed a convergent triangulation mixed methods design. Both quantitative and qualitative data were collected. The Yukawa Mentorship Evaluation Tool was used to measure program effectiveness. Descriptive analyses were conducted by a member of the research team. Data was coded by two members of the research team and findings were audited for consistency.

**Results:** A total of 51 participants (27 mentors, 24 mentees) completed the survey, and 13 (7 mentors, 6 mentees) participated in semi-structured interviews. Five themes emerged: Mentorship Characteristics, Program Administration, Perceived Program Benefits, Barriers to Mentorship, and Recommendations for Improvement.

**Conclusion:** The BPC mentorship program represents a historic step toward addressing unmet needs of Black medical residents in Canada. Participants expressed high satisfaction and highlighted areas for improvement. Racially concordant mentorship was seen as particularly valuable in addressing unique challenges faced by Black learners. The findings from this study provide critical insights into best practices for future mentorship programs, advancing diversity and equity in medicine while supporting Black learners’ success.

## Introduction

Despite making up 4.3% of the Canadian population and 16.1% of the Racialized population, Black people constitute only 1.5% of the Canadian physician workforce^1^. While the Black population is projected to double by 2041^1^, the disproportionately low numbers of Black physicians highlight significant barriers within the medical profession. Black learners face emotional abuse, isolation, and high attrition rates during training, underscoring the need for systemic change^2–4^.

Racial categorization historically served to justify colonialism and slavery, with modern racial identities reflecting entrenched social hierarchies^5^. In medical schools, ostensibly race-neutral structures continue to uphold these inequities, as racial bias influences admissions, curriculum design, and support systems^6^. Addressing racism in medicine requires dismantling these structures that sustain it, and actively building new structures to replace it^6^. Increasing representation within medical education exposes learners to different cultures and experiences that prepares them to meet the needs of the population they serve ^7^ and having patient-provider race concordance improves survival and reduces mortality in Black population^8–10^. While cultural competency training has helped reduce racial disparities in healthcare^11^, racially concordant mentorship is said to be particularly effective in supporting Black medical trainees^4^. Irrespective of the well documented positive impacts on patient outcomes, the need for our medical training environments to be free from racism should never require justification.

Globally, mentorship has proven critical during high-stress periods, such as through career progression, rigorous training, and academic challenges. A report conducted by the Higher Education Funding Council of England^12^ highlighted mentorship and peer support as key to success across all higher education sectors. Within medicine specifically there is evidence of the positive impact of mentorship on participant satisfaction, and on objective outcomes such as promotion and retention ^13–15^. In our medical training system which, after pre-clerkship, remains primarily apprenticeship-based^16^, mentors serve to help socialize and navigate our complex academy.

Rooted in ancient traditions of guidance and protection^17^,mentorship is particularly crucial in addressing racial disparities within medical training. Echoing the proverb “it takes a village to raise a child”, strong support systems are viewed as vital for success in Black and African communities. This encompasses support not only from parents but also from peers and elders, ensuring the sharing of wisdom and guidance across generations. Mentorship programs specifically designed for underrepresented minorities in medicine (URiM) have improved academic and professional outcomes, provided more equitable training opportunities, fostered career progression and acts as a protective factor against racial differentials ^15,18–20^. Given the cultural emphasis on community support in Black communities, one might assume Black learners have robust access to mentorship. However, many report barriers to accessing mentorship, with a few studies noting participants who reported lacking access to quality mentorship entirely ^4,21,22^.

In the Scarborough Charter^23^, 58 Canadian higher education institutions publicly committed to advancing Black inclusion and excellence in higher education in Canada. A key directive of this charter is the development of mentorship programs for Black students and professionals, which are structured to facilitate growth through support from mentors with similar backgrounds. The charter emphasizes institutional accountability, with regular assessments and transparent reporting on the success of these initiatives in promoting Black inclusion ^23^.

### Black Physicians of Canada Mentorship Program

Recognizing a gap in racially concordant mentorship for Black residents, fellows, and early-career physicians, Black Physicians of Canada (BPC) and the Black Residents Physicians of Ontario (BRPO) conducted a Canada-wide needs assessment in 2020, which found that nearly 80% of Black residents lacked formal mentorship and desired guidance from mentors of similar racial and professional backgrounds ^21^. In response, BPC established the BPC mentorship program in July 2021, a pioneering initiative in Canada, designed to support Black medical learners and early career physicians through race-concordant mentorship. This program was developed to address the specific needs of Black physicians, offering individual mentorship, proactive support, and tailored communal education sessions to foster community and belonging. Mentors and mentees were provided with a handbook outlining ground rules, which included 4 mandatory meetings throughout the year. Program-wide events over the year covered topics such as transition to residency, dealing with microaggressions and the hidden curriculum, and finances. Through mentors with similar racial backgrounds and lived experiences, Black residents, fellows, and early-career physicians gained unique insights and guidance often absent in traditional mentorship programs.

This study explores the experiences of mentors and mentees in the first iteration of the program, identifying perceived facilitators and barriers to mentorship. The evaluation provides valuable insights into the specific mentoring needs of Black residents and early career physicians, highlighting the unique benefits of racially concordant mentorship programs in medical training. Drawing on our findings, we make recommendations to guide the development and future evaluation of similar programs, shaping what “effective” mentorship is and helping guide similar programs to success.

## Materials and Methods

### Study Design

This study employed a convergent triangulation mixed methods design ^24^ to examine the effectiveness of the BPC mentorship program. Quantitative survey data and qualitative semi-structured interview data were collected concurrently. The surveys gathered data on socio-demographics and self-reported effectiveness of the mentorship program using the Mentorship Evaluation Tool (MET) ^25^. Semi-structured interviews were conducted with a subset of mentors and mentees to explore their lived experiences in the BPC mentorship program. Our study design enabled mentorship to be explored from different perspectives, which were then brought together in a convergent mixed-methods approach to enhance our understanding of the mentorship experience ^26^. All study procedures were approved by the Research Ethics Board at the University of Toronto.

### Study Setting and Participants

The BPC mentorship program had 58 mentor-mentee pairs in the program’s first year, comprising Black residents and early-career physicians matched by gender and specialty to a mentor. All 116 program participants in this cohort were invited via email to participate in the study. Survey respondents were subsequently invited for interviews, with all interviewees completing the survey beforehand and receiving an honorarium for their participation.

### Data Collection and measures

Questions for the survey and semi-structured interviews were developed based on responses from a needs assessment survey ^21^ and consultations with experts in mentorship, equity, medical education, qualitative research (CY, UN, CK, MN, MTB, RS, AM). The survey comprised of 19 questions for mentors and 21 for mentees, assessing mentorship effectiveness using a seven-point Likert scale (1 = Strongly Disagree to 7 = Strongly Agree) based on the Yukawa Mentor Evaluation Tool (MET) ^25^. The interview guide was tailored for mentors and mentees, addressing themes such as incentives, mentorship experiences, pairing process, facilitators and barriers to mentorship, mentorship impact, and race-concordant mentorship’s role in medicine. After obtaining informed consent, semi-structured interviews were conducted over Zoom between May 2023 and August 2023, with sessions averaging 45 minutes for mentees and 50 minutes for mentors. All interviews were audio-recorded, transcribed verbatim, and de-identified before data entry into NVivo 12.7.0 for qualitative analysis.

### Data Analysis

Descriptive analyses were conducted by a member of the research team (MT) on the responses to closed survey questions. Interview data were analysed using an inductive thematic analysis approach ^27^. The inductive thematic approach involves a six-step approach: familiarising with data, generating initial codes, searching for themes, reviewing themes, defining and naming themes, and finally producing the report. Two research team members (CO and NC) independently coded the data, with discrepancies resolved through discussions with a multidisciplinary team, and findings were subsequently audited for consistency.

## Results

Overall, 51 BPC program participants (27 mentors and 24 mentees) **(Table 1)** completed the survey, for an overall response rate of 44%. Semi-structured interviews were held with thirteen participants (seven mentors and six mentees).

**Table 1.** Demographic characteristics of Black mentors and mentees who completed surveys and interviews to explore their experiences in a race-concordant mentorship program.

| Characteristics | Mentors |  | Mentees |  |
| --- | --- | --- | --- | --- |
|  | *n* | % | *n* | % |
| Gender |  |  |  |  |
| Male | 14 | 52 | 8 | 33 |
| Female | 13 | 48 | 16 | 67 |
| Age Range |  |  |  |  |
| 20-29 | 0 | 0 | 2 | 8 |
| 30-39 | 4 | 15 | 15 | 63 |
| 40-49 | 13 | 48 | 7 | 29 |
| 50-59 | 9 | 33 | 0 | 0 |
| >60 | 1 | 4 | 0 | 0 |
| First Language |  |  |  |  |
| English | 24 | 89 | 22 | 92 |
| French | 3 | 11 | 1 | 4 |
| Other | 0 | 0 | 1 | 4 |
| Continent/Region of Origin |  |  |  |  |
| Africa | 8 | 30 | 11 | 46 |
| North America | 15 | 56 | 10 | 42 |
| Europe | 0 | 0 | 1 | 4 |
| Caribbean | 3 | 11 | 2 | 8 |
| Middle East | 1 | 4 | 0 | 0 |
| Specialty |  |  |  |  |
| Medicine | 6 | 22 | 8 | 33 |
| Surgery | 3 | 11 | 5 | 21 |
| Family medicine | 10 | 37 | 7 | 29 |
| Psychiatry | 1 | 4 | 1 | 4 |
| Pediatrics | 2 | 7 | 0 | 0 |
| Radiology | 1 | 2 | 2 | 8 |
| Other | 4 | 15 | 1 | 4 |
| Medical School |  |  |  |  |
| Dalhousie University Faculty of<br>Medicine | 3 | 15 | 1 | 4 |
| McGill University Faculty of<br>Medicine | 1 | 5 | 0 | 0 |
| McMaster University Michael G.<br>DeGroote School of Medicine | 2 | 10 | 0 | 0 |
| Queen's University School of<br>Medicine | 0 | 0 | 1 | 4 |
| University of Alberta Faculty of | 0 | 0 | 1 | 4 |
| Medicine and Dentistry |  |  |  |  |
| University of British Columbia | 2 | 10 | 1 | 4 |
| Faculty of Medicine |  |  |  |  |
| University of Calgary Cumming | 2 | 10 | 2 | 8 |
| School of Medicine |  |  |  |  |
| University of Manitoba Max Rady | 1 | 5 | 1 | 4 |
| College of Medicine |  |  |  |  |
| University of Ottawa Faculty of | 1 | 5 | 6 | 25 |
| Medicine |  |  |  |  |
| University of Toronto Faculty of | 5 | 25 | 8 | 33 |
| Medicine |  |  |  |  |
| Université de Montréal Faculté de | 1 | 5 | 1 | 4 |
| Médecine |  |  |  |  |
| Université de Sherbrooke Faculté | 1 | 5 | 0 | 0 |
| de Médecine et des Sciences de la |  |  |  |  |
| Santé |  |  |  |  |
| Western University Schulich | 1 | 5 | 2 | 8 |
| School of Medicine and Dentistry |  |  |  |  |
| Training level |  |  |  |  |
| Resident | 0 | 0 | 13 | 54 |
| Fellow | 0 | 0 | 1 | 4 |
| Practice Setting |  |  |  |  |
| Academic physician | 16 | 62 | 5 | 21 |
| Community Physician | 10 | 39 | 5 | 21 |

Our findings can be summarized in five domains: (1) Mentorship Characteristics (2) Program Administration (3) Perceived Program Benefits (4) Barriers to mentorship and (5) Recommendations for Improvement. The content in each domain is summarised below, using descriptive statistics from surveys and anonymized quotes from interviews to illustrate the concepts.

### 1) Mentorship characteristics

Mentees valued honest feedback, expertise, and a growth mindset, while mentors emphasized active listening. Both groups highlighted relationship quality and feedback as top indicators of successful mentorship. Shared racial and gender identities strengthened connections, with mentees finding essential guidance on navigating microaggressions and the emotional impact of racism. Drawing from their own experiences, mentors offered vital support, fostering a sense of belonging and confidence in mentees.

In survey responses, mentees highlighted: providing honest and constructive feedback (96.3%); sharing relevant knowledge and expertise (92.6%); and having a growth mindset (85.2%), as important factors. While mentors generally agreed on these most important characteristics, they also highlighted exhibiting active listening skills (90.3%). When asked what metrics should be measured to evaluate “good” mentorship, “relationship quality” was rated highest as a measure of good mentorship by both mentees (100%) and mentors (93.6%). (81.5%) of mentees and (80.1%) of mentors rated “quality of feedback received” as the second most important measure of good mentorship.

The significance of shared identity, particularly in terms of race and gender, emerged as an important factor in fostering a meaningful mentorship relationship amongst all participants. For example, one mentee described “*The city I trained in didn’t have very many people of colour or Black female physicians, and so I was excited about the idea of having a Black woman as a mentor. She was very helpful as someone who has three kids so she had been through residency and being pregnant.*”

Black mentors with similar lived experiences played a pivotal role by providing valuable resources, offering guidance during challenging rotations, and fostering a sense of belonging that transcended the formal boundaries of mentorship. For example, one mentee describes the one valuable encounter with their mentor “*I found her extremely helpful when I felt like I wasn’t treated very well on a rotation. She had a similar experience. She told me about what she had felt she wished she had done differently. So, I was able to take those experiences that she had and was willing to share with me, and make it so that I had a better kind of resolution*.”

Several mentors who have themselves benefitted from mentorship or have overcome systemic career discrimination saw this mentorship program as a way to pay forward their knowledge and experience. As one mentor reflected, *“There is an intrinsic benefit in terms of giving back and seeing someone else succeed and you feel like you’ve hopefully played a small role in building their confidence.”*

### 2) Program administration

Regular communication via email and reminders kept mentors and mentees engaged, while the thoughtful pairing process matched participants with shared interests, creating stronger connections. High event attendance highlighted participants’ appreciation, despite some challenges with virtual interactions. Overall, participants were satisfied with the program’s supportive structure and clear guidance.

According to survey responses, approximately 71% of mentees attended BPC mentorship events over the program year, and approximately 76% of the mentees cited the usefulness of the mentorship events. While 84% of the mentors attended the BPC mentorship events over the program year, while 76% “agreed” or “strongly agreed” that the events were useful. Although in interview responses, participants shared the challenges posed by virtual platforms in fostering meaningful mentorship connections within the BPC program, suggesting the preference for *“more tangible and impactful in-person interactions.”*

The manual mentor-mentee pairing process by the program organizers was hailed as a thoughtful and impactful endeavor, with mentees expressing gratitude for being matched in subspecialties that were not only professionally relevant but also personally resonant. Participants expressed that the manual matching helped ease the typical challenge of participants coordinating matching themselves, it ensured adequate follow-up and provided guidelines on roles. For example, one mentor described how *“the program was well organized, in terms of matching us and making sure that we understood our expectations, and I found that this program was great.”*

### 3) Perceived program benefits

The BPC mentorship program fostered a supportive environment where mentors and mentees connected through shared cultural experiences, creating a foundation of trust and openness. This program allowed participants to discuss both career-related and personal challenges, including those often unique to Black professionals in medicine, such as navigating microaggressions and balancing well-being with professional demands. Many participants found this holistic approach highly valuable, as it addressed aspects of their professional journeys that were rarely acknowledged in other mentorship programs.

In survey responses, mentors highlighted “review of career goals” (80.1%), “building coping skills, teamwork and collaborative skills” (57.7%) and “help navigate difficult circumstances such as microaggression” (42.4%) are most helpful to mentees during the program. While mentees highlighted “job prospects and recommendations” (91.7%), “sponsorship/networking” (79.2%) and “psychosocial support from a fellow Black physician” (79.2%) and “help to navigate microaggressions” (75.0%) as most important in their lives. Overall, 95% of the mentees rated their overall satisfaction with the mentoring program positively, and no participants disagreed with the statement.

Those who had prior experience with mentorship, compared their non-Black and Black mentors and felt in the BPC program, it was easier and safer bringing up certain conversations in their mentor-mentee relationship. For example, one mentor commented that “*Even though there’s still diversity even within a group of Black people, there is also a lot more in common. So it was easier to break the ice, it was easier to start off the relationship*.”

A recurrent theme was appreciating the holistic nature of the program, extending beyond career guidance to encompass the broader spectrum of life and academia. Many mentees commented that their mentors were impactful in providing guidance on issues such as *“dealing with microaggressions”*, *“addressing the emotional toll of racism”*, and *“navigating the complexities of a medical career.”* As one mentee explained: *“What BPC has a unique take on is the aspect of wellness from a minority person perspective and inclusion is definitely what is the key to success of this program.”* While another mentee described “…*with the BPC program, I’m able to discuss both career, life, and academic stuff, and it’s more holistic. I feel I don’t have to worry too much in the background about what I’m saying, who I’m saying it to and what the implications are on my career.”*

Some participants described their mentorship program experience as not only professionally enriching but also *“personally affirming”*, and expressed an *“increased sense of belonging”* and *“increased confidence.”* As one mentee shared: *“I think it has positively affected me in that I now feel a little bit less of the imposter syndrome”*.

Participants shared how financial constraints significantly impacted the learning experience of Black medical learners and highlighted the impact of the mentorship program on exploring topics such as “*financial discussions around how to prepare for the financial burden of residency*”, “*when transitioning to work, how to negotiate that contract*” and “*how to have a career trajectory planned*” and how these discussions added a practical dimension to mentorship relationships.

Some participants felt there was a lack of support from their institutions in providing access to mentorship and overall guidance on dealing with negative experiences. For example, some mentees shared that they had no previous formal mentorship prior to the BPC program, *“I didn’t have access to mentorship in the department that I was in, so the BPC program was an opportunity to get a mentorship with an additional benefit of someone who’s had these challenges that people who have given me advice to up until this point did not have to face.”* Another mentee described “*I think that the biggest contribution to the mentorship was that my mentor really introduced me to the vast range and variety of career arrangements that are possible within the field.*”

### 4) Barriers to mentorship

Key barriers included the demanding time commitment for mentors, the limitations of virtual communication for distant mentor-mentee pairs, and the complex pressures of navigating racial issues within the medical field.

Participants highlighted the Black tax experienced by Black physicians. A mentor expressed that “*One of the biggest challenges was the significant time commitment expected from participants, often compounding the challenges of an already demanding profession.”* Some participants commented on the impact of geographical distance on the effectiveness of the mentorship. As a nationwide program some mentor-mentee pairs were in different cities so had to rely on online virtual methods of communication for the duration of the program. For example, one mentee described *“I would say the geography was the only thing that didn’t work for me and having to do things virtually.”* Participants shared the challenges posed by virtual platforms in fostering meaningful mentorship connections within the BPC program.

Many participants commented on the nuanced challenges faced by Black individuals pursuing careers in medicine. For example, one mentor commented that “*with Black mentees, they have the added element of the emotional toll of racism that impacts the wellness perspective*” while another mentor described “*I think all levels need discussion on how race interferes or helps their progress. For the non-Black mentees, this does not usually come up. They’re not really thinking about barriers in this way. They may only be thinking about the competitive nature, for example of CARMS but they’re not necessarily worrying about who’s going to evaluate me for the next block?*”

### 5) Recommendations for improvement

Looking toward future iterations of the BPC mentorship program, this section explores the importance of continuous support and engagement as well as recommendations suggested by participants.

Mentors expressed a desire to have more tailored and specific continuing professional development and guidance on Racialized and Black mentorship techniques. For example, one mentor commented that they would “*like to have vignettes and videos resources that recreate navigating different mentorship scenarios, that will be valued.”* While another mentor recommended *“printed information, like a booklet or brochure with information about topics that would come up, to give ideas of what to reflect on, things mentees might want to discuss and what sort of things in your experience you want to kind of take a deep dive into.”*

Participants expressed that networking with allies would be a valuable resource for mentorship, particularly extending networking beyond the immediate community. As a mentor described “*A lot of our students and learners do end up finding mentee mentorship through other allies. So networking is key, and not just being focused on our community, but looking at a wider pool of skills and talent and willingness.*”

Participants also made recommendations for improving the mentor-mentee matching. Some participants suggested the incorporation of mentees’ long-term career goals into the matching process, to ensure a more tailored and effective mentorship relationship. As one mentee explains “*One question that I think would be great is asking the mentees or prospective mentees where they see their career going in the next five to ten years. And then matching the mentees’ five-to-ten-year plan to mentors so that somebody could help them get to that goal.*”

## Discussion

In response to the growing recognition of the need for improved mentorship and training experiences for underrepresented (UR) students in fields like medicine^21,22,28^, the launch of the Black Physicians of Canada’s mentorship program marks a critical step toward addressing the challenges and the need for mentorship among Black residents in Canada. Through understanding the roles and functions of mentorship and the importance of shared identity in mentorship, we can better support Black medical trainees.

Our study findings demonstrate that race-concordant mentorship is perceived as especially important by participants as it alleviates the need to explain the context and nuances of their lived experiences, creating an environment of safety and trust ^29^. Similar to previous research, our participants described race-concordant mentorship as desirable as it addresses the unique needs of racial minorities, including psychosocial support, financial difficulties, social guidance, and emotional toll of racism ^30–32^. Consistent with previous research, our mentees reported that having mentors of similar racial or ethnic backgrounds helped them envision their own potential and increased confidence in their future careers^29^. Our results align with previous studies that emphasise mentorship as a crucial component in career development and improved training experience^33^.

Our study highlights key elements of effective mentoring relationships that can guide medical education leaders and stakeholders in designing impactful mentoring programs for their trainees. Our participants described mentor characteristics, mentor-mentee relationship quality and feedback as critical factors for successful mentoring. Having a mentor who actively listens, participates in the development and training of the mentee was an important factor that increased satisfaction among the participants. An additional important factor was the facilitation of dyad matching by program organizers rather than having mentees to choose their mentor themselves as this reduced the burden of coordinating an “ideal match” prior to initiating a mentoring relationship. Our participants emphasized that mentoring should be formalized and well organized like any other activity in academic medicine^34^. Optimal mentor-mentee relationships can be facilitated by fostering professional and social networking opportunities^35^. Findings from the study also revealed that beyond race concordance, gender and medical specialty concordance is perceived as especially important in increasing compatibility between mentor-mentee pairs.

Our recommendations to all academic stakeholders and the public for implementing race-concordant mentorship program in medical education include:

### 1. Black Self-Determination and Centralized Black-Led Mentorship

The growing number of Black medical students and residents highlights the urgent need for more Black physician mentors. While programs like the BPC mentorship program and the Black Physicians Association of Ontario’s mentorship pods have made progress in increasing the pool of mentos available to Black learners, the demand for mentors outpaces supply. The underrepresentation of Black physicians in Canada’s medical workforce, limits the pool of mentors available to Black learners.

To mitigate this gap, we recommend centralized mentorship networks, group mentorship models, and recognizing mentors through stipends, awards, or academic incentives. Hiring more Black faculty and strengthening support systems for Black learners are essential to fostering diversity, inclusion, and a sustainable mentorship pipeline in medicine.

### 2. Development of evaluation frameworks and mentorship models specific to the Black experience

As we work towards creating safer learning environments and increasing mentorship opportunities for Black learners, it is crucial to develop mentorship strategies that specifically address their unique needs. Mentorship frameworks must be tailored to foster empowerment, resilience, and a sense of belonging, ensuring that Black students thrive academically and personally.

Building communities of practice is another key element for the ongoing development of Black medical learners. Programs like the Community of Support at the University of Toronto (number) have been successful in creating a collaborative environment for supporting underrepresented undergraduate students pursuing medicine, but similar initiatives are lacking at the postgraduate level. These communities will provide opportunities for mentors to refine their skills, while allies will learn how to better support Black learners. This collaborative approach will strengthen mentorship, and create inclusive, supportive learning environments for Black learners, ultimately advancing diversity in medicine.

Mentorship programs must adopt trauma-informed, strength-based approaches that celebrate cultural heritage and address systemic challenges. Robust evaluation frameworks are also needed to measure psychosocial outcomes, such as belonging and community, alongside academic and career progression.

### 3. Real investments and long-term commitments to supporting Black excellence and learner safety

To support Black medical learners, institutions must establish formalized spaces for connection, community-building, and shared experiences. These spaces combat isolation, enhance belonging, and improve well-being. Feedback highlights the critical role of psychological safety, which is often lacking in existing mentorship programs. Race-concordant mentorship, where mentors share cultural or racial backgrounds with learners, fosters understanding, support, and reduces imposter syndrome. Without safe spaces, Black learners face additional challenges navigating medical education. Creating these environments not only prioritizes mental health but also facilitates networking and mentorship, empowering learners to overcome unique obstacles. By fostering community and psychological safety, institutions can ensure Black learners thrive in their medical training.

The study has several limitations. Our sample only included Black physicians in the BPC program, who may differ from other residents, fellows, and staff physicians not in the program. Therefore, our results may not generalize to all Black residents, fellows, and staff physicians. However, given that this was a Canada wide program with participants from 13 medical schools across the country, they represent a diverse sample group. The use of virtual interviews and surveys, though necessary due to logistical constraints, may have limited rapport and depth, with in-person interactions potentially allowing richer discussion. An additional limitation is that survey and interview participation were voluntary, and there may be potential for responder bias. However, through a mixed methods approach this study provides further insights on the nuanced experiences of Black physicians in Canada.

## Conclusion

As more Black students enter medical institutions, it’s essential to provide support that promotes their retention and career success. Medicine’s historical under representation has created systemic barriers for marginalized groups. Resources like mentorship from physicians with shared identities and experiences offer Black students the community and guidance necessary for advancement. These study results will serve to inform best practices in race concordant mentorship programs within medical education, tailored to address the unique challenges faced by Black medical learners. An area of further study would be to determine the long-term effects of race-concordant mentoring on academic, clinical and professional performance.

In conclusion, we draw on Black feminist visionary bell hooks^36^ on the power of humanism and trauma-informed pedagogy. Like patients, our learners thrive when seen as multidimensional human beings, carrying the resilience of our ancestors. By drawing on the authority of experience in our communities we transgress colonial educational practices, and education becomes the practice of freedom.

## Data Availability

All relevant data are within the manuscript and its Supporting Information files.

## Acknowledgements and Disclosures

Acknowledgements

The authors thank The Black Physicians of Canada (BPC) Research Group, Hadal El-Hadi MD and Menal Huroy MD, all of whom provided valuable insight and constructive feedback on this project at various stages of development.

## Ethical Approval

All study procedures were approved by the Research Ethics Board at the University of Toronto.

## Previous presentations

The findings of this study have been previously presented in part at the 2024 International Congress for Academic Medicine (ICAM).

